# World Health Organization Danger Signs to predict bacterial sepsis in newborns: A pragmatic prospective cohort study

**DOI:** 10.1101/2023.05.09.23289739

**Authors:** Omolabake Akinseye, Constantin R. Popescu, Msandeni Chiume-Kayuni, Michael A. Irvine, Norman Lufesi, Tisungane Mvalo, Niranjan Kissoon, Matthew O. Wiens, Pascal M. Lavoie

## Abstract

**Background:** The World Health Organization (WHO) has developed danger signs (DS) to help front-line health workers triage interventions in children with severe illnesses. Our objective was to evaluate the extent to which DS predict bacterial sepsis in young infants presenting with acute illness.

**Methodology/Principal Findings:** This prospective study evaluated nine DS in infants younger than 3 months with suspected sepsis in a large regional hospital in Lilongwe, Malawi, between June 2018 and April 2020. The main outcomes were positive blood or cerebrospinal fluid (CSF) cultures and mortality. Blood (n=85/401) and CSF (n=2/204) cultures were positive in 21.2% and 1% of infants, respectively (N=401; gestational age mean ± SD: 37.1±3.3 weeks, birth weight 2865±785 grams). In-hospital deaths occurred in 9.7% (N=39/401) of infants (61.5% within 48h of admission). In univariate analyses, all DS were associated with mortality except for temperature instability and tachypnea, whereas “infant was unable to feed” was the only DS significantly associated with bacterial sepsis.

After co-variable adjustments, number of DS predicted mortality (OR: 1.75; 95%CI: 1.43–2.16; p<0.001; AUC-ROC: 0.756) but not positive cultures (OR 1.08; 95%CI: 0.92–1.30; p=0.336). Whether potential bacterial contaminants were included or not did not change results meaningfully.

**Conclusion/Significance:** DS predicted fatal outcomes but not positive cultures in a large regional hospital setting. These data imply that the incidence of bacterial sepsis and attributable mortality are unlikely to be accurate based on clinical signs alone, in infants in LMIC settings.

## Introduction

Three million children die each year worldwide in their first month of age, with a significant proportion of these deaths occurring during the first week(1). Severe infections (including sepsis, pneumonia, diarrhea and viral diseases) are common causes of serious illnesses in young infants (2). Up to 25% of cases in low- and middle-income countries (LMICs) have identified bacterial causes (3). However, the lack of diagnostic resources, particularly in LMICs makes it difficult to accurately quantify this burden. This gap has enormous implications in terms of implementing policies and guidelines for the use of antimicrobials to prevent these deaths (1). At the individual level, identifying bacterial causes for sepsis is crucial to prevent fatalities and guide the correct empiric antibiotic therapy in the context of rising antimicrobial resistance. Indeed, the misuse, or even the overuse, of antibiotics carries a risk of subsequent treatment failure due to colonization by resistant bacteria (4, 5).

To assist front-line health workers, the World Health Organization (WHO) and United Nations International Children’s Emergency Fund developed algorithms in the early 1990s to help triage interventions for sick children (6). This Integrated Management of Childhood Illness (IMCI) strategy included the identification of clinical signs to alert the need for escalation of care, later termed Danger Signs (DS) (6). Subsequently, the Young Infant Clinical Signs Study Group carried out a multi-centered study in six LMICs and found that seven signs (i.e., history of difficulty feeding, grunting, convulsions, movement only when stimulated, respiratory rate ≥60 breaths per minute (bpm), severe chest indrawing, temperature below 35.5°C or above 37.5°C, prolonged capillary refill, cyanosis and stiff limbs) best predicted the need for higher level of care, with a sensitivity of 74% to 85%, and specificity of 75% to 79%, depending on infants’ age (i.e., 0 to 6 days versus 7 to 59 days old) (7). A systematic review of five studies on 17,506 infants found similar results (8).

Over time, the use of DS has been extended to provide a reasonable basis for the initiation of empiric antibiotic treatment (9). However, these signs have neither been developed nor validated to diagnose sepsis of bacterial etiology. Neonatal disorders such as transient tachypnea of the newborn, hypoglycemia, hyaline membrane disease, birth asphyxia, and viral infections (to mention only a few), present in a fashion similar to bacterial sepsis (10). Hence, a key question is whether DS can reasonably identify newborns or young infants who truly benefit from antibiotic treatment. Only a few studies have attempted to answer this question, but certainly none that have, to the best of our knowledge, been conducted prospectively, in a pragmatic setting where clinicians, not research staff, pragmatically reported DS (11).

The objective of this study was to prospectively determine the extent to which WHO DS predict bacterial sepsis in young infants, in a LMIC hospital setting where front-line health workers were asked to assess and record the presence of DS in infants who presented with suspected sepsis.

## Materials and Methods

### Study design, setting & population

This study was a pragmatic cohort study, and secondary analysis of data from a parent study that aimed to identify molecular signatures of bacterial sepsis in young infants with suspected sepsis (12). In the parent study, infants below 3 months of age were sequentially enrolled “around the clock” at the time they presented with suspected sepsis before, to the neonatal or pediatric units at Kamuzu Central Hospital in Lilongwe, Malawi between June 5^th^, 2018, and April 6^th^, 2020. To minimize bias towards false-negative blood culture, infants were excluded if they had received antibiotics for >4 hours at the time of enrollment. In the original (parent) study, about 851 infants presented with suspected sepsis and the vast majority were not enrolled because staff were unable to get consent -either parents declined or the conditions were not sufficiently permissive for proper consent due to other clinical demands. Enrolled infants were followed during hospitalization.

The Kamuzu Central Hospital the largest referral hospital in central Malawi, where approximatively 3,000 infants are admitted to its neonatal unit annually. “Suspected sepsis” was defined as per the clinical staff, essentially in an infant that looked unwell where bacterial sepsis could not be excluded clinically. Before the study, the staff at Kamuzu Central Hospital did not have routinely access to blood culture or other ancillary tests, IV fluids and gavage feeds were also most often not available due to limited resources and supplies. Available interventions include antibiotics, assistance with feeding and routine surgeries when necessary. Oxygen and continuous positive pressure ventilation were only available to an extremely limited number of infants, on a limited basis when the hospital had electrical power. Invasive mechanical ventilation was not possible. Written informed consent was obtained from all parents/guardian of each participant, in English or Chichewa (main local language). During the study, the authors did not have access to information that could identify participants, with the exception of the consent forms who were kept under the responsibility of the local study PI (MCK). The study was approved by the University of British Columbia’s Children’s and Women’s Research Ethics Board (#H16-02639) and the National Health Science Research Committee of Malawi (#17/8/1819).

### Clinical data and blood samples collection

Each infant had an initial clinical evaluation by a healthcare worker (or staff, either a nurse or clinical officer), and the presence of the following DS were prospectively collected using electronic forms, on a tablet device: history of poor feeding (interviewing the parent/caregiver), entered as “feeding well”, “unable to feed” or “uninterested in feeding”, respiratory rate (visually counted by attended clinician), axillary body temperature (measured using an electronic Welch Allyn SureTemp thermometer, model 692), drowsiness/difficulty to wake, grunting, severe chest recession, central cyanosis and convulsions (defined pragmatically by attending clinician). All infants also had a standardized aerobic blood culture (minimum 2 mL of blood) collected within 4 hours of presentation, using a rigorously implemented aseptic protocol (with in-training of all front-line healthcare providers prior to study initiation), as detailed (13). Infants also had cerebrospinal fluid (CSF) and urine cultures collected as clinically indicated. Fast breathing was defined as a respiratory rate of ≥60 bpm. Temperature instability was defined as a body temperature <35.5°C or >38°C. Blood cultures were processed into BACTEC PEDS Plus culture bottles incubated in a BD BACTEC 9050 instrument.In keeping with a pragmatic design, the study did not attempt to enforce strict criteria to define DS or illness status. Identification of organisms from blood or CSF cultures was completed in-house using bioMerieux API, and BD Crystal kits at a local University of North Carolina-affiliated research facility, as described (12). Culture results were made available to clinicians within 5 days. Bacterial isolates that could be restored (∼60% of isolates) were also analyzed for further confirmation of identity (not shown) on a Microflex LT MALDI-TOF mass spectrometer, using the BioTyper software version 3.1 (Bruker Daltonik, Germany).

### Outcome measures

The primary outcomes of interest were *mortality*, and *culture-positive sepsis* defined as the bacterial growth of a known neonatal pathogen in blood or CSF culture sample. Micro-organisms not commonly considered as pathogens in term or near-term infants, such as coagulase-negative Staphylococcus, Micrococcus or Bacillus were excluded (14), but these potential contaminants were included in a sensitivity analysis using an alternate bacterial sepsis outcome (**Supplemental Table 1**). In-hospital mortality was recorded until discharge home or transfer to another hospital facility. Data were considered missing when infants were transferred within 72 hours after initial admission to the hospital.

### Statistical Analysis

In the parent study, the sample size was set conveniently at 500 infants over 2 years, but the study needed to be stopped in April 2020 due to the COVID-19 pandemic. Demographic characteristics were reported descriptively using the mean/median with dispersion. The following nine WHO DS were used as predictors: *not feeding well, convulsions, drowsy/difficult to wake, movements only when stimulated or no movements, fast breathing, grunting, severe chest recessions, temperature instability (hypo/hyperthermia), central cyanosis*. All quantitative variables were analyzed as such, except for fast breathing and temperature instability which were categorized according to WHO definitions (see above). Odds ratios were calculated for each DS and the main outcomes of bacterial sepsis or mortality. Logistic regressions were conducted between the number of DS and the main outcomes. In separate analyses, odds ratios were calculated using logistic regressions between mean DS (i.e., the number of DS divided by the available DS data) to determine the effect of missing DS data. Unadjusted logistic regression models, including demographic variables, were conducted to determine the predictive value of the DS. The predictive performance of DS for each outcome was assessed using the area under receiver operator characteristic curve (AUC-ROC). Sociodemographic variables associated with mortality, as well as their independent effects were analyzed using regression models. Only complete cases were used in the analysis. Statistical significance was considered at p-value <0.05 with no adjustment made for multiple testing in the unadjusted models. Analyses were performed using R (version 4.1.0) and SPSS (version 20; IBM, Armonk, NY, USA).

## Results

### Study population

All infants originally enrolled in the parent study were included, for a total of 401 infants enrolled in the original study and included in this analysis. The majority were term or late preterm infants, born vaginally, presented from home in their first week of post-natal life, and had roughly equal female/male distribution (**Table 1**). Most infants were reported by clinicians to be ill at the time of initial presentation (69.3%; 278/401), 22.2% (89/401) were severely ill, and 8.5% appeared well (34/401). In total, 79.8% (320/401) presented at least one DS. Overall, 56.9% (228/401) of infants had been ill for at least 24 hours prior to initial presentation, and their parents had traveled a median distance of 17 kilometers, often by foot or taxi, to reach the nearest hospital. In total, 94/401 (23.4%) infants had received antibiotics prior to the blood culture, for a median of 2 hours (range 0.75 to 3.5 hours). The duration of antibiotic use in infants ranged from 1 to 30 days (**Table 1**). Few infants with negative blood culture had antibiotics discontinued after 48 hours (21/401, 5.2%).

**Table 1:** Socio-demographic characteristics of the study population.

| Participant characteristics |  | N with data |
| --- | --- | --- |
| Gestational age at birth (weeks), mean (SD) | 37.1 (3.3) | 397 |
| Birth weight (gram), mean (SD) | 2865 (785) | 380 |
| Female, no (%) | 185 (46) | 401 |
| Admitted from, no (%) |  | 401 |
| Inborn | 161 (40) |  |
| Transferred from another facility | 224 (56) |  |
| Home | 16 (4) |  |
| Post-natal age at presentation (days), median (IQR) | 4 (2-16) | 401 |
| Mode of delivery, no (%) |  | 398 |
| Cesarean section | 101 (25.2) |  |
| Vaginal delivery | 297 (71.8) |  |
| Duration of illness before presentation to hospital (hours), median (IQR) | 24 (12-48) | 341 |
| Distance of travel to hospital (km), median (IQR) | 17 (11-25) | 342 |
| Illness status, number on infants (%) |  | 401 |
| Well | 34 (8.5) |  |
| Ill | 278 (69.3) |  |
| Severely ill | 89 (22.2) |  |
| Duration of antibiotics (days), median (IQR) | 6 (5-9) | 401 |
| Duration of hospital stay (days), median (IQR) | 7 (5-9) | 401 |
| Danger Signs, number of infants (%) |  |  |
| Not feeding well | 171 (43.3) | 395 |
| Convulsions | 28 (7.0) | 401 |
| Drowsy / Difficult to wake | 33 (8.2) | 401 |
| Movements only when stimulated or no movements | 26 (6.5) | 401 |
| Fast breathing $\geq 60$ per min | 104 (27.1) | 384 |
| Grunting | 30 (7.5) | 401 |
| Severe chest recessions | 51 (12.7) | 401 |
| Temperature instability $>38^{\circ}\text{C}$ or $<35.5^{\circ}\text{C}$ | 118 (32.7) | 361 |
| Central cyanosis | 29 (7.2) | 401 |

### Outcome data

Blood (n = 85/401) and CSF (n = 2/204) cultures were positive in 21.2% and 1% of the infants, respectively. However, about half (46 / 85) had bacterial types considered to be contaminants (e.g., *bacillus, micrococcus, corynebacterium* and *coagulase-negative staphylococcus* species and non-pathogenic *streptococcus* species such as *oralis*), so the actual proportion with bacterial sepsis, considering only known pathogens in term and near-term infants, was effectively 9.7% (39/401).

Excluding contaminants, gram-positive bacteria (53.8%; 21/39) were slightly more prominent than gram-negative bacteria (46.2%; 18/39) (**Supplemental Table 1**). Among all infants with presumed sepsis, pathogen sepsis was more common in the inborn (17/161) than in the infants transferred from another hospital facility or from home (22/240) (p=0.029). The proportion of gram-positive sepsis was comparable between inborn infants (10/240) versus in infants transferred from another hospital or home (11/161) (p=0.240). Three infants were transferred before 72 hours, so among infants in whom data were available, the mortality was 9.7% (39/398), of which 61.5% (24/39) died within 48h.

### Associations with mortality and culture-positive outcomes

The following DS were significantly associated with mortality (**Table 2**): *not feeding well, convulsions, drowsy/difficult to wake, movements only when stimulated or no movements, grunting, severe chest recessions, central cyanosis*. In contrast, *fast breathing* or *temperature instability* were not significantly associated with mortality. The cumulative number of DS was also significantly associated with mortality (OR 1.75 [95%CI: 1.43 – 2.16] per additional DS; p<0.001) (**Supplemental Figure 1**). Admission weight, but not age of the mother, gestational age, birth weight and sex were significantly associated with mortality (**Supplemental Table 2**). When adjusting for all of these co-variables, cumulative number of DS remained significantly associated with mortality (**Supplemental Table 2**; p<0.001).

**Table 2:** Associations between DS and mortality.

| Variable | Odds ratio (OR) | 95%CI | P value |
| --- | --- | --- | --- |
| Not feeding well | 7.21 | (3.27 - 18.2) | <0.001 |
| Convulsions | 3.52 | (1.31 - 8.59) | 0.008 |
| Drowsy / Difficult to wake | 5.04 | (2.12 - 11.4) | <0.001 |
| Movements only when stimulated or no movements | 4.89 | (1.88 - 11.9) | <0.001 |
| Fast breathing $\geq 60$ per min | 1.86 | (0.87 - 3.85) | 0.100 |
| Grunting | 3.95 | (1.54 - 9.34) | 0.002 |
| Severe chest recessions | 3.01 | (1.34 - 6.37) | 0.005 |
| Temperature instability $>38^{\circ}\text{C}$ or $<35.5^{\circ}\text{C}$ | 1.70 | (0.75 - 3.75) | 0.191 |
| Central cyanosis | 4.15 | (1.62 - 9.87) | 0.002 |

Bacterial sepsis with a known pathogen was significantly associated with mortality (OR 3.29 [95%CI: 1.37 – 7.36]; p = 0.005). However, the only DS significantly associated with bacterial sepsis when considering only known neonatal pathogens was *not feeding well* (OR 2.04 [95%CI: 1.05 – 4.06]; p=0.038) (**Table 3**). Associations between convulsions, drowsiness, lethargy (i.e., movements only when stimulated or no movement), tachypnea, grunting, severe chest recession, temperature instability and central cyanosis were all non-significant (**Table 3**). Association was between cumulative number of DS and bacterial sepsis was also non-significant (OR 1.21 [95% CI: 0.98 – 1.47]; p=0.072) in regression models (**Supplemental Figure 2**). At discharge, clinical staff selected non-bacterial sepsis in 76.3% (306/401), bacterial sepsis in 15.5% (62/401) and “other non-sepsis syndromes” in 8.2% (33/401) of cases, from these options, for study purposes.

**Table 3:** Association between DS and bacterial sepsis when organism is a known pathogen.

| Variable | Odds ratio (OR) | 95%CI | P value |
| --- | --- | --- | --- |
| Not feeding well | 2.04 | (1.05 - 4.06) | 0.038 |
| Convulsions | 0.69 | (0.11 - 2.45) | 0.626 |
| Drowsy / Difficult to wake | 0.91 | (0.21 - 2.73) | 0.886 |
| Movements only when stimulated or no movements | 1.75 | (0.49 - 4.90) | 0.327 |
| Fast breathing $\geq 60$ per min | 1.16 | (0.53 - 2.38) | 0.703 |
| Grunting | 2.54 | (0.89 - 6.31) | 0.058 |
| Severe chest recessions | 1.61 | (0.62 - 3.71) | 0.288 |
| Temperature instability $>38^{\circ}\text{C}$ or $<35.5^{\circ}\text{C}$ | 1.80 | (0.86 - 3.70) | 0.113 |
| Central cyanosis | 0.67 | (0.11 - 2.34) | 0.588 |

When including all positive blood cultures (**Supplemental Table 3**), culture positivity was not significantly associated with mortality (OR 1.77 [95% CI: 0.83 – 3.6]; p=0.123), or any DS. The cumulative number of DS was also not significantly associated with bacterial sepsis when including positive cultures with potential bacterial contaminants (OR 1.21 [95% CI: 0.98 – 1.47]; p=0.0715).

## Discussion

This study found that most WHO DS were significantly associated with mortality, but only one DS (i.e., not feeding well) as significantly associated with culture-proven bacterial sepsis. Additionally, the majority of DS were not associated with culture-proven bacterial sepsis in this LMIC neonatal cohort. The results held true whether only bacterial pathogens, or potential bacterial contaminants were considered. To the best of our knowledge, the current study is the first to pragmatically examine whether DS predict bacterial sepsis in infants. These findings were obtained prospectively in an LMIC setting where DS were identified by front line healthcare workers as they would normally do pragmatically outside a research protocol – we assumed that this study design would more closely reflects the real-life utility of DS. Few studies have attempted to determine how DS can predict bacterial sepsis caused by bacterial pathogens in newborns or young infants. A retrospective Brazilian study reviewed data on 83 sick neonates over a 12 months period, and came to the same conclusion that DS alone or in combination did not predict severe serious bacterial infection (15). The findings of the current study are also consistent with those of another study carried out in a primary care, rural setting in Bangladesh, India and Pakistan. In the latter study, clinical signs could not differentiate infants with viral or bacterial illnesses (16). The current study findings have implications for a hospital setting, but further research is needed to determine the generalizability of the results to other hospital contexts.

In addition to the main findings, the current study also found that only a minor proportion of infants with suspected sepsis showed positive blood culture. Only few other studies have prospectively estimated the incidence of bacterial sepsis in LMIC cohorts, and it is therefore useful to present the current findings in the context of those other studies. A hospital centre study in Delhi, India, included 13,530 neonates (all comers; mean GA±SD: 36.0±3.4 weeks) monitored daily for clinical signs of sepsis with per-protocol completion of a sepsis work-up (blood culture and lumbar puncture) prior to initiating antibiotics, and found a 14.3% incidence of culture-positive sepsis (17), which is similar to the findings of the current study. Another study in community settings in Madagascar, Senegal and Cambodia reported 8.2% culture-confirmed sepsis among 514 infants assessed for “possible serious bacterial infection” out of 3,688 neonates (18). One large study followed 12,622 live-births up to 60 days of age in rural India between April 2002 and March 2005, with daily home visits and clinical assessment, and standardized blood and CSF cultures following enrollment of infants with suspected sepsis. It reported a 6.3% incidence of culture-confirmed sepsis (53 with positive blood cultures out of 842 infants assessed, excluding potential contaminants), for a population-level incidence of culture-confirmed sepsis of 6.7 per 1,000 births (19). Taken together, these findings suggest that bacterial sepsis may be uncommon in this age group even in LMICs, when sepsis is clinically suspected.

Blood cultures are the gold standard for diagnosing bacterial sepsis in young infants (20). However, these tests lack sensitivity in neonates (21). In the current study we were careful to prescribe a minimum blood volume to maximize culture yield. Yet, a key question is whether we misestimate the proportion of bacterial sepsis in infants in whom cultures are negative? The Aetiology of Neonatal Infection in South Asia (ANISA) study (22) also attempted to identify the cause of sepsis for 6022 cases of possible serious bacterial infections in infants in the first 60 days after birth, in a population of 63,114 livebirth). Bacterial and viral (e.g., respiratory syncytial virus) causes were identified in 16% and 12% of cases, respectively. For the remaining 72% cases, no pathogen could be identified despite systematic blood and respiratory cultures, which is consistent with findings from the current study.

Evidently the use of antibiotics in the current study was high, with the vast majority of infants receiving antibiotics for a minimum of five days during hospitalization, as per hospital practices. As in other LMIC settings, blood cultures were not routinely available at the study site prior to the study. As such, no approach had been developed to stop antibiotics after 48 hours given a negative blood culture. In another study in Thailand, the duration of antibiotic use was also high with durations of 15 and 8 days for culture-proven and culture-negative early-onset neonatal sepsis, respectively (23). These results highlight the need to rely on DS, and potential coercive effect of the lack of laboratory resources capable of providing basic laboratory test results to help “rule out” sepsis (24).

A strength of this study is its prospective design, with standardized blood culture procedures to ensure that the chances of contamination during blood sampling were minimized, and robust microbiological identification methods including clinical laboratory protocols on-site augmented by advanced MALDI-TOF species determination in another, North American clinical microbiology laboratory. The strong association between bacterial sepsis and mortality suggest that the infants with positive cultures were correctly identified. However, a number of study limitations should be acknowledged. Although a sizeable proportion of the recruited infants presented from home, data were collected in a single hospital-type setting; as such, caution should be exercised when generalizing to other hospital settings, or extrapolating to non-hospital primary care settings. For logistical reasons CSF was collected from only approximately half of the infants, which may have underestimated the incidence of bacterial sepsis. Moreover, due to the extremely limited diagnostic capabilities of the study LMIC settings, in most cases, causes of deaths or alternate diagnoses could not be determined. Similarly, the study did not include non-bacterial testing, which precluded identification of viral sepsis syndromes.

In conclusion, our study validates the use of WHO DS to predict fatal outcomes, but not bacterial sepsis, in young infants. This study further emphasizes the need for better diagnostics for bacterial sepsis, in order to ensure that health gains continue to improve, in areas of the world where sepsis represents a huge burden and where antimicrobial resistance is also increasing at alarming rates (25).

## Data Availability

We are happy to make the data available but will need to check with our REB to obtain appropriate permissions.

## Abbreviations

WHO: World Health Organization
DS: danger signs
LMICs: low and middle-income countries
CSF: cerebrospinal fluid
AUC-ROC: Area Under Curve – Receiver Operator Characteristics
bpm: breaths per minute.

## Acknowledgments

We thank Dr. David Goldfarb for expert input into data interpretation and Amber Johnson from the British Columbia Children’s Hospital Microbiology Laboratory for confirming bacterial isolates by mass spectrometry.

